# High orientation and low delayed recall in the standardisation of the Spanish version of the Montreal Cognitive Assessment (MoCA) in elders of Peru

**DOI:** 10.1101/2024.06.16.24308987

**Authors:** Lucia Bulgarelli, Emilia Gyr, Jose Villanueva, Koni Mejía, Claudia Mejía, Renato Paredes, Sheyla Blumen

## Abstract

**INTRODUCTION:** The elderly in Peru face healthcare barriers in detecting cognitive impairment and dementia due to a lack of validated tools. The Montreal Cognitive Assessment (MoCA) effectively detects early dementia, assessing visuo-spatial function, executive function, naming, memory, attention, language, abstraction, and orientation.

**METHODS:** This study aims to standardise the Spanish version of the MoCA for the elderly in Lima. The test was administered to 338 elders from three institutions: Municipality of San Miguel District, San José Obrero Polyclinic, and EDMECON. Regression-based normed scores were computed, adjusted for age and education.

**RESULTS:** Our results show high orientation scores and low delayed recall, highlighting cognitive strengths and weaknesses in our sample. Age and education significantly influenced cognitive performance, with education as the strongest predictor.

**DISCUSSION:** This study offers normative data for the Peruvian elderly, aiding the clinical use of MoCA in Peru. We discuss appropriate cut-off points and cultural sensitivity in the Peruvian context.

## 1 INTRODUCTION

Dementia represents a set of pathologies whose common characteristic is the death of neuronal cells, triggering a degenerative and irreversible process affecting the nervous system ^1,2^. Dementia is a clinical syndrome that involves irreversible loss of cognitive functions, especially memory, significantly compromising daily activities ^3^. Diagnosis requires a substantial decrease in patient autonomy and quality of life compared to a premorbid stage, not attributable to other medical or psychiatric causes ^4^. Neurocognitive disorders are among the most prevalent age-related disorders worldwide and lead to substantial morbidity and mortality ^5–7^.

The prevalence of dementia among individuals aged 60 years and older is approximately 8.5% in Latin America ^8^ with projections indicating an increase from 7.8 million in 2013 to 27 million by 2050^9,10^. The Global Burden of Disease (GBD) study shows that neurodegenerative diseases in Peru, measured in Disability Adjusted Life Years (DALY), rose from the fifth to the third largest contributor to neurological diseases ^11^. A recent study in Peru ^12,13^ found a 6.85% prevalence of dementia in 1,532 individuals over 65 in Lima, with Alzheimer’s disease (AD) as the predominant diagnosis. Plausible factors contributing to these findings include the ageing of the population, poor access to health services, low diagnostic capacity of dementia in primary care services, and limited recognition of symptoms by family members and caregivers ^13^. Given the recent increase in life expectancy, it is plausible that AD incidence will rise, potentially tripling by 2050^4,14^.

There is no cure for dementia, only drugs to slow its progression ^15^. For this reason, early diagnosis is beneficial for patients and their caregivers and can result in substantial cost savings for healthcare systems ^16^. In fact, early diagnosis provides numerous benefits to the patient: (a) greater and more effective adherence to pharmacological and rehabilitative treatments, (b) the possibility of participating in treatments that are still experimental, and (3) the possibility of organising one’s life in relation to the progression of the disease ^17^.

Diagnosing dementia in Latin America faces critical issues: tests are rarely performed in primary care. This leads to a high number of cases of dementia that go unnoticed and misdiagnosed ^18^. In this context healthcare workers can identify dementia with a 66% positive predictive value after a few hours of training ^18^.

Several cognitive screening tools for MCI and dementia ^19,20^ have low sensitivity for early detection ^21^. Most tests are complex, needing trained personnel and specialised equipment ^22^, often unavailable in primary care. Consequently, MCI is frequently unrecognised in primary care ^22^.

Promoting research on short cognitive tests validated for Peru ^23^ is essential, similar to neuropsychological test validation projects in other countries (for example, the NEURONORMA projects in Spain ^24^ and Colombia ^25^). To date, several short cognitive tests have been validated in the context of Lima (Peru), including the Clock Drawing Test (CDT) ^26 27^; the Addenbrooke’s Cognitive Examination (ACE) ^28^; the INECO frontal screening test (IFS) ^29^ and the memory impairment test (M@T) ^30,31^. However, the MoCA test (“https://mocacognition.com/”) has not been validated in the Peruvian context.

MoCA is presented as a brief, reliable, and valid test for neurocognitive evaluation given its high degree of sensitivity and specificity for MCI and dementia ^22,32^. Currently, the MoCA is available in multiple translated versions, including adaptations for individuals with visual impairments, as well as video and telephone formats. These adaptations improve the accessibility and usability of the test, particularly in longitudinal studies, by minimising sample bias and mitigating participant attrition ^33^. Thus, it arises as a promising alternative for the detection of early cognitive impairment by general practitioners ^34^. In the Latin America region, there is a tendency to use the original Spanish version of the MoCA test due to the scarcity of studies related to its validity and cross-cultural adaptation. This situation may result in practices that are not in compliance with international diagnostic standards ^35^. The objective of this study is to standardise the Spanish version of the Montreal Cognitive Assessment Test (MoCA) for the elderly in Lima-Peru, assessing the impact of age, sex, and educational level. In addition, the norms for the Lima elderly population will be developed taking into account age and educational level.

## 2 METHODS

### 2.1 Participants

Participants were recruited from three institutions within the Metropolitan Lima: the EDMECON medical centre, the San José Obrero Polyclinic, and the San Miguel Municipality. The sample consisted of 338 seniors over 60 years of age (122 male, 216 female), including both native Spanish speakers and those who spoke Spanish as a second language (with proficiency exceeding 10 years). Sociodemographic data, including variables such as sex, age, date of birth, years of education, and medical diagnoses, were systematically collected (Table 1). The age was divided into six ranges: 60-64, 65-69, 70-74, 75-79, and 80+ years after the research conducted by the National Institute of Informatics and Statistics (INEI) of Peru ^36^. Similarly, years of education were divided into three ranges: 0-6, 7-11 and 12 + years following the education levels reported by INEI ^37^: Primary, Secondary, and Higher.

**TABLE 1.** Sociodemographic characteristics of the sample.

|  |  | Male (n=122) |  | Female (n =216) |  | Total(n=338) |  | X<sup>2</sup>(p) |
| --- | --- | --- | --- | --- | --- | --- | --- | --- |
|  |  | n | % | n | % | n | % |  |
| Age group | 60 - 64 years | 11 | 9.0 | 40 | 18.5 | 51 | 15.0 | 8.0(0.08) |
|  | 65 - 69 years | 26 | 21.3 | 44 | 20.3 | 70 | 20.7 |  |
|  | 70 - 74 years | 24 | 19.6 | 49 | 22.6 | 73 | 21.5 |  |
|  | 75 - 79 years | 22 | 18.0 | 35 | 16.2 | 57 | 16.8 |  |
|  | >80 years | 39 | 31.9 | 48 | 22.2 | 87 | 25.7 |  |
| Educational level | Primary (0 - 6 years) | 19 | 15.5 | 50 | 23.1 | 69 | 20.4 | 3.3(0.18) |
|  | Secondary (7 - 11 years) | 51 | 41.8 | 90 | 41.6 | 141 | 41.7 |  |
|  | Higher (>12 years) | 52 | 42.6 | 76 | 35.1 | 128 | 37.8 |  |

Individuals with auditory or visual impairments, psychiatric diagnoses, concurrent cerebrovascular pathologies, intellectual disability, traumatic brain injuries, and substance abuse disorders were excluded from the sample.

### 2.2 Instrument

Montreal Cognitive Assessment (MoCA) is a validated cognitive screening tool renowned for its efficacy in the early detection of dementia ^38^. The administration of the test typically takes around 10 minutes. The test evaluates eight key cognitive domains: visuospatial and executive function, naming, memory, attention, language, abstraction, and orientation (see Table 2). The original MoCA scoring system incorporates a correction factor, adding one point to the total score for individuals with fewer than 12 years of education (28). Here, we aim at estimating the adjustments due to age and education for our population.

**TABLE 2.** MoCA Item Distribution and Assigned Scores.

| Cognitive domain | Subtest | Task | Maximum score |
| --- | --- | --- | --- |
| Visuospatial Skills / Executive Functions | Shortened TMT-B | It is evaluated using a graphic alternation task adapted from the Trail Making Test B. | 1 |
|  | Cube Copying | It is evaluated with the copy of a geometric cube. | 1 |
|  | Clock Drawing Test | It is evaluated using the copy of the clock test. | 3 |
| Naming | Identifying Animal Figures (Lion, Rhino, Camel) | It is evaluated using three items, naming by visual confrontation of three animals with a low degree of familiarity. | 3 |
| Attention | Digits | Direct reverse digits | 1 |
|  | Inverse digits | Reverse digits | 1 |
|  | Tapping Test - letter A | Sustained attention | 1 |
|  | Subtraction of 7 from 100 | A series of subtractions | 3 |
| Language | Phrase Repetition | Repetition of two complex sentences | 2 |
|  | Word Fluency | Phonetic fluency (number of words that begin with the letter F during one minute) | 1 |
| Abstraction | Similarities | It is made up of two abstract verbal reasoning items | 2 |
| Memory | Delayed Recall | It consists of two learning trials of five words (they do not score) for which questions are asked in a deferred manner after five minutes. | 5 |
| Orientation | Day of the Month (Date) | Temporal and spatial orientation is evaluated. | 1 |
|  | Month |  | 1 |
|  | Year |  | 1 |
|  | Day of the week |  | 1 |
|  | Place |  | 1 |
|  | Location |  | 1 |

### 2.3 Procedure

Contact with participants was conducted within the context of a medical appointment at the three institutions. In the case of the Private Centre Medical EDMECON and the National Polyclinic of San José Obrero, the evaluation was performed as part of the patient’s medical evaluation, and authorisation was requested to obtain the data by informed consent. In the case of the Municipality, the data collection was parallel to the outpatient medical care provided by the institution, where elderly were visited at their homes.

### 2.4 Data Analysis

We examined the potential effects of age, years of education, and sex in the raw scores of MoCA for the total sample. We evaluated sex differences in the total score and each of the cognitive domains of the MoCA using the Mann-Whitney U tests. We used the Bonferroni ^39^ correction of *p*-values to control for the family-wise error rate of multiple comparisons.

We explored associations of age and years of education with the total score and each of the cognitive domains of the MoCA using the Kendall correlation coefficient (*τb*) and the determination coefficient (*r*2). Here, we employed the Bonferroni correction to control for the family-wise error rate for each test. Potential differences in cognitive performance according to age group and educational level were analysed using the Kruskal-Wallis H test and Bonferroni corrected post hoc Dunn tests ^40^.

Finally, we computed regression-based normed scores of the total MoCA scores adjusted for age and years of education following the procedure described in the NEURONORMA project ^24,41^. First, raw scores were converted to percentile ranks (Pc). These ranks were rescaled to a truncated normal distribution (M: 10, SD: 3) bounded between 2 and 18 to generate Scaled Scores adjusted for age (*SSa*). This procedure was used for the entire sample and each age group.

Next, adjustments for years of education were modelled using a linear regression:

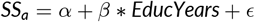

The estimated regression coefficient (*β*) here was employed to calculate Scaled Scores adjusted for age and years of education (*SSae*) ^42^:

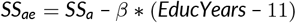

We selected a cut-off point of 11 years of education because it represents the period of basic education (Primary and Secondary) in Peru. Data preprocessing, visualisations and statistical analyses were performed with Pandas ^43^, Seaborn ^44^ and Pingouin ^45^ software libraries of the Python programming language.

## 3 RESULTS

The sociodemographic characteristics of the 338 participants in terms of age, education level, and sex are shown in Table 2. The mean cognitive performance of the total sample was 18.2 ± 5.6 for the total MoCA score. The scaled scores for each subtest are shown in Figure 1.

**FIGURE 1.**
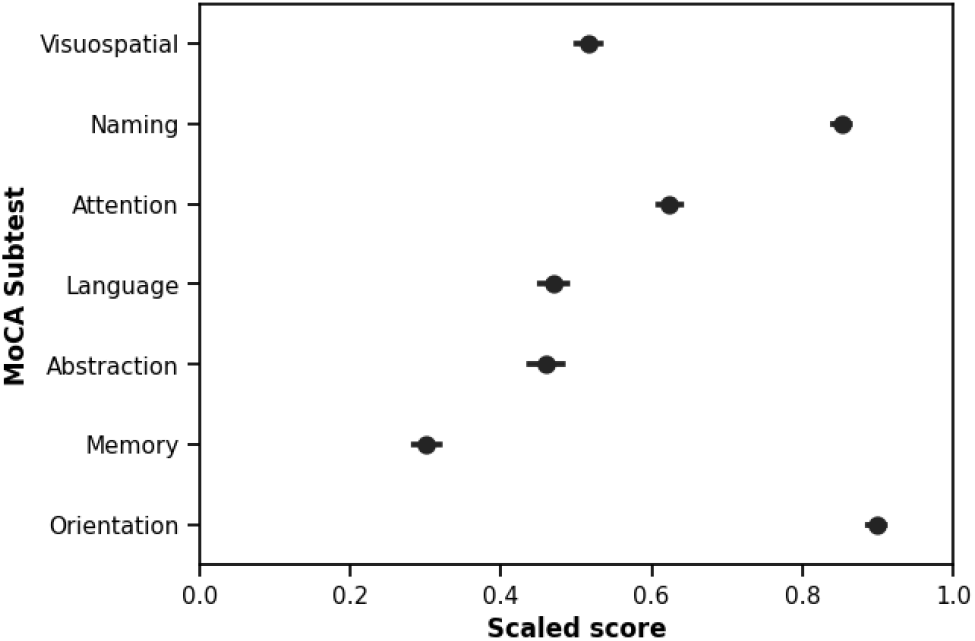
Scaled scores of MoCA subtests.

### 3.1 Effects of sex, age and education

We did not find significant differences in the total scores of MoCA grouped by sex. However, we observed that men scored higher on identification (U = 10984.5, *p* = .023) and attention (U = 10540.5, *p* = .015) than women. No significant differences were found for other cognitive domains (see Table 3).

**TABLE 3.**
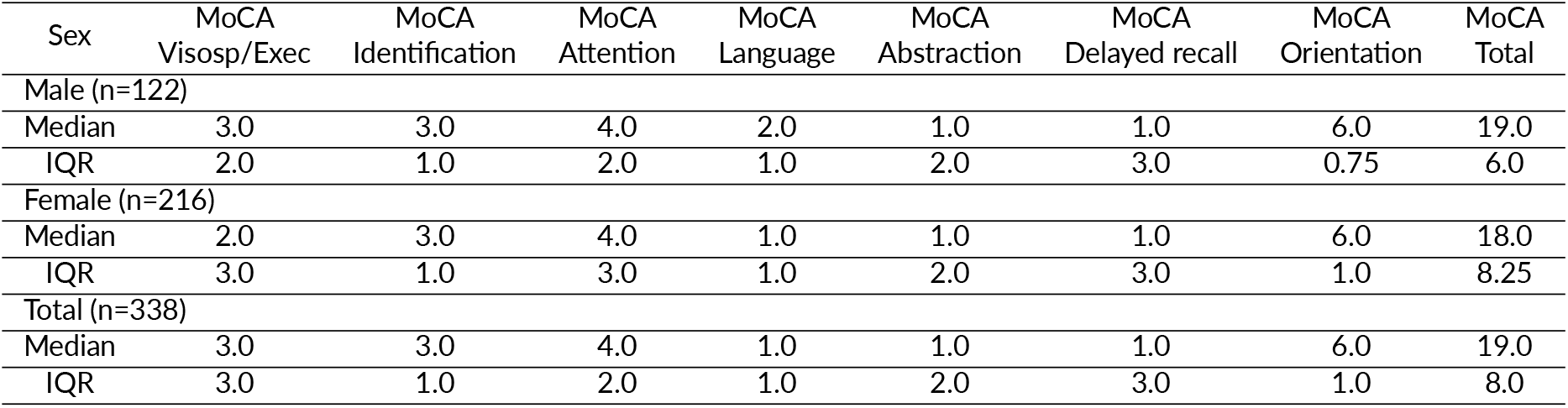
MoCA scores grouped by sex.

| Sex | MoCA<br>Visosp/Exec | MoCA<br>Identification | MoCA<br>Attention | MoCA<br>Language | MoCA<br>Abstraction | MoCA<br>Delayed recall | MoCA<br>Orientation | MoCA<br>Total |
| --- | --- | --- | --- | --- | --- | --- | --- | --- |
| Male (n=122) |  |  |  |  |  |  |  |  |
| Median | 3.0 | 3.0 | 4.0 | 2.0 | 1.0 | 1.0 | 6.0 | 19.0 |
| IQR | 2.0 | 1.0 | 2.0 | 1.0 | 2.0 | 3.0 | 0.75 | 6.0 |
| Female (n=216) |  |  |  |  |  |  |  |  |
| Median | 2.0 | 3.0 | 4.0 | 1.0 | 1.0 | 1.0 | 6.0 | 18.0 |
| IQR | 3.0 | 1.0 | 3.0 | 1.0 | 2.0 | 3.0 | 1.0 | 8.25 |
| Total (n=338) |  |  |  |  |  |  |  |  |
| Median | 3.0 | 3.0 | 4.0 | 1.0 | 1.0 | 1.0 | 6.0 | 19.0 |
| IQR | 3.0 | 1.0 | 2.0 | 1.0 | 2.0 | 3.0 | 1.0 | 8.0 |

The results of the correlation analysis for age and years of education are shown in Table 4. Both age and years of education were significantly correlated with the MoCA total score and its subtests. We found that age was a significant negative predictor of the total MoCA score and its subtests. In contrast, we found that years of education were a significant positive predictor of the total score of MoCA and its subtests. Age and years of education explain 12.17% and 24.67% of the total scores of MoCA, respectively.

**TABLE 4.** Correlation analysis between the Montreal Cognitive Assessment (MoCA) and the demographic variables of age and years of education.

|  | Age |  | Years of Education |  |
| --- | --- | --- | --- | --- |
| | $\tau_b$ | $r^2$ | $\tau_b$ | $r^2$ |
| Visospatial/Executive | -0.22* | 0.09* | 0.34* | 0.16* |
| Identification | -0.17* | 0.04* | 0.19* | 0.06* |
| Attention | -0.15* | 0.04* | 0.27* | 0.12* |
| Language | -0.16* | 0.04* | 0.34* | 0.17* |
| Abstraction | -0.19* | 0.06* | 0.31* | 0.13* |
| Delayed recall | -0.17* | 0.05* | 0.18* | 0.05* |
| Orientation | -0.20* | 0.05* | 0.27* | 0.11* |
| MoCA Total | -0.24* | 0.12* | 0.37* | 0.24* |

\*p < 0.05

We found a significant main effect of the age group on the total MoCA scores (H = 40, *p* < .001). Post hoc pairwise comparisons revealed significant differences between the 80 + (Me = 15, IQR = 7) group and the 60-64 (Me = 22, IQR = 6.5) (*p* < .001), 65-69 (Me = 21, IQR = 8.75) (*p* < .001) and 70-74 (Me = 19, IQR = 6) (*p* = .004) groups. Similarly, significant differences were found between the 60-64 group and the 75-79 (Me = 19, IQR = 10) group (*p* = .031).

Similarly, we found a significant main effect of the educational level on the total scores of MoCA (H = 79.80, *p* < .001). Post hoc pairwise comparisons revealed significant differences between the Primary education group (Me = 13, IQR = 6) and the Secondary (Me = 19, IQR = 5) (*p* < .001) and the Higher Education groups (Me = 22, IQR = 7) (*p* < .001). Furthermore, significant differences were found between the Secondary and Higher Education groups (*p* < .001).

### 3.2 Age and education adjusted norms

Age-adjusted scale scores obtained from our sample are presented in Table 5. Total scores, percentile values (Pc), age-adjusted scaled scores (*SSa*), and scores for the total sample and the five age groups are presented. Following ^42^, we suggest cutoff scores to classify cognitive performance in the test based on the following criteria:

**TABLE 5.**
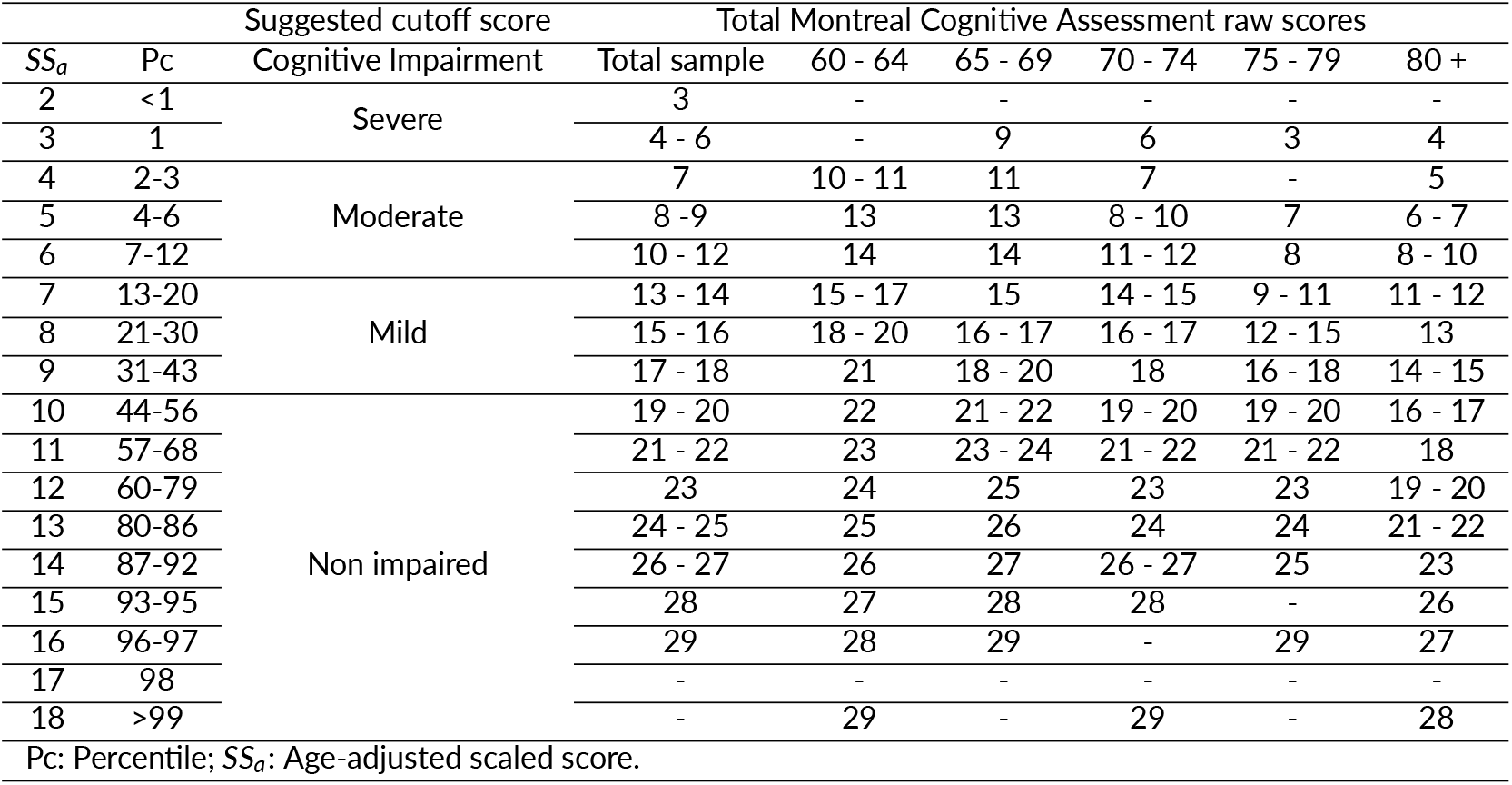
Scaled scores and percentiles for each age group.

- Obtaining a scaled score of 10 or more and a Pc of 44-56 (or more) would not be considered impairment.
- Obtaining a scaled score of 7 (one standard deviation below the mean) and a Pc of 13-43 would correspond to mild cognitive impairment.
- Obtaining a scaled score of 4 (two standard deviations below the mean) and a Pc of 2-12 would correspond to moderate cognitive impairment.
- Obtaining a scaled score of 2 (three standard deviations below the mean) and a Pc < 1 would correspond to severe cognitive impairment.

For the total sample, cognitive performance is considered normal or non-impaired when a score of ≥ 17 is obtained.

Years of education significantly predicted *SSa* (*r*^2^ = .239, *p* < .001) with a coefficient of 0.347. Following ^24^, we used this coefficient to calculate age and education adjusted scaled scores (*SSae*) for each combination of *SSa* and years of education observed in the sample. We present *SSae* in Table 6.

**TABLE 6.** Scales scores adjusted for age and years of education.

| SS<sub>a</sub> | Pc | Years of education |  |  |  |  |  |  |  |  |  |  |  |  |  |  |  |  |  |  |  |  |
| --- | --- | --- | --- | --- | --- | --- | --- | --- | --- | --- | --- | --- | --- | --- | --- | --- | --- | --- | --- | --- | --- | --- |
|  |  | 0 | 1 | 2 | 3 | 4 | 5 | 6 | 7 | 8 | 9 | 10 | 11 | 12 | 13 | 14 | 15 | 16 | 17 | 18 | 19 | 20 |
| 2 | <1 | 5.0 | 5.0 | 5.0 | 4.0 | 4.0 | 4.0 | 3.0 | 3.0 | 3.0 | 2.0 | 2.0 | 2.0 | 1.0 | 1.0 | 0.0 | 0.0 | 0.0 | -1.0 | -1.0 | -1.0 | -2.0 |
| 3 | 1 | 6.0 | 6.0 | 6.0 | 5.0 | 5.0 | 5.0 | 4.0 | 4.0 | 4.0 | 3.0 | 3.0 | 3.0 | 2.0 | 2.0 | 1.0 | 1.0 | 1.0 | 0.0 | 0.0 | 0.0 | -1.0 |
| 4 | 2-3 | 7.0 | 7.0 | 7.0 | 6.0 | 6.0 | 6.0 | 5.0 | 5.0 | 5.0 | 4.0 | 4.0 | 4.0 | 3.0 | 3.0 | 2.0 | 2.0 | 2.0 | 1.0 | 1.0 | 1.0 | 0.0 |
| 5 | 4-6 | 8.0 | 8.0 | 8.0 | 7.0 | 7.0 | 7.0 | 6.0 | 6.0 | 6.0 | 5.0 | 5.0 | 5.0 | 4.0 | 4.0 | 3.0 | 3.0 | 3.0 | 2.0 | 2.0 | 2.0 | 1.0 |
| 6 | 7-12 | 9.0 | 9.0 | 9.0 | 8.0 | 8.0 | 8.0 | 7.0 | 7.0 | 7.0 | 6.0 | 6.0 | 6.0 | 5.0 | 5.0 | 4.0 | 4.0 | 4.0 | 3.0 | 3.0 | 3.0 | 2.0 |
| 7 | 13-20 | 10.0 | 10.0 | 10.0 | 9.0 | 9.0 | 9.0 | 8.0 | 8.0 | 8.0 | 7.0 | 7.0 | 7.0 | 6.0 | 6.0 | 5.0 | 5.0 | 5.0 | 4.0 | 4.0 | 4.0 | 3.0 |
| 8 | 21-30 | 11.0 | 11.0 | 11.0 | 10.0 | 10.0 | 10.0 | 9.0 | 9.0 | 9.0 | 8.0 | 8.0 | 8.0 | 7.0 | 7.0 | 6.0 | 6.0 | 6.0 | 5.0 | 5.0 | 5.0 | 4.0 |
| 9 | 31-43 | 12.0 | 12.0 | 12.0 | 11.0 | 11.0 | 11.0 | 10.0 | 10.0 | 10.0 | 9.0 | 9.0 | 9.0 | 8.0 | 8.0 | 7.0 | 7.0 | 7.0 | 6.0 | 6.0 | 6.0 | 5.0 |
| 10 | 44-56 | 13.0 | 13.0 | 13.0 | 12.0 | 12.0 | 12.0 | 11.0 | 11.0 | 11.0 | 10.0 | 10.0 | 10.0 | 9.0 | 9.0 | 8.0 | 8.0 | 8.0 | 7.0 | 7.0 | 7.0 | 6.0 |
| 11 | 57-68 | 14.0 | 14.0 | 14.0 | 13.0 | 13.0 | 13.0 | 12.0 | 12.0 | 12.0 | 11.0 | 11.0 | 11.0 | 10.0 | 10.0 | 9.0 | 9.0 | 9.0 | 8.0 | 8.0 | 8.0 | 7.0 |
| 12 | 60-79 | 15.0 | 15.0 | 15.0 | 14.0 | 14.0 | 14.0 | 13.0 | 13.0 | 13.0 | 12.0 | 12.0 | 12.0 | 11.0 | 11.0 | 10.0 | 10.0 | 10.0 | 9.0 | 9.0 | 9.0 | 8.0 |
| 13 | 80-86 | 16.0 | 16.0 | 16.0 | 15.0 | 15.0 | 15.0 | 14.0 | 14.0 | 14.0 | 13.0 | 13.0 | 13.0 | 12.0 | 12.0 | 11.0 | 11.0 | 11.0 | 10.0 | 10.0 | 10.0 | 9.0 |
| 14 | 87-92 | 17.0 | 17.0 | 17.0 | 16.0 | 16.0 | 16.0 | 15.0 | 15.0 | 15.0 | 14.0 | 14.0 | 14.0 | 13.0 | 13.0 | 12.0 | 12.0 | 12.0 | 11.0 | 11.0 | 11.0 | 10.0 |
| 15 | 93-95 | 18.0 | 18.0 | 18.0 | 17.0 | 17.0 | 17.0 | 16.0 | 16.0 | 16.0 | 15.0 | 15.0 | 15.0 | 14.0 | 14.0 | 13.0 | 13.0 | 13.0 | 12.0 | 12.0 | 12.0 | 11.0 |
| 16 | 96-97 | 19.0 | 19.0 | 19.0 | 18.0 | 18.0 | 18.0 | 17.0 | 17.0 | 17.0 | 16.0 | 16.0 | 16.0 | 15.0 | 15.0 | 14.0 | 14.0 | 14.0 | 13.0 | 13.0 | 13.0 | 12.0 |
| 17 | 98 | 20.0 | 20.0 | 20.0 | 19.0 | 19.0 | 19.0 | 18.0 | 18.0 | 18.0 | 17.0 | 17.0 | 17.0 | 16.0 | 16.0 | 15.0 | 15.0 | 15.0 | 14.0 | 14.0 | 14.0 | 13.0 |
| 18 | >99 | 21.0 | 21.0 | 21.0 | 20.0 | 20.0 | 20.0 | 19.0 | 19.0 | 19.0 | 18.0 | 18.0 | 18.0 | 17.0 | 17.0 | 16.0 | 16.0 | 16.0 | 15.0 | 15.0 | 15.0 | 14.0 |
| Pc: Percentile; SS<sub>a</sub>: Age-adjusted scaled score. |  |  |  |  |  |  |  |  |  |  |  |  |  |  |  |  |  |  |  |  |  |  |

Pc: Percentile; SS<sub>a</sub>: Age-adjusted scaled score.

## 4 DISCUSSION

The purpose of this study was to standardise the Spanish version of the Montreal Cognitive Assessment (MoCA) for the elderly population in Lima. Standardising this test in Peru is crucial for an accurate cognitive evaluation, considering the high prevalence of neurocognitive disorders ^8,13^ and the lack of cognitive tools adapted to the sociodemographic and cultural context of the country. Furthermore, early detection of cognitive disorders such as dementia improves patient prognosis, quality of life, and reduces economic and social burdens ^3^, which is critical for developing countries.

We examined the effect of sex, age, and education on MoCA scores. In line with previous research using MoCA ^24,42^, we found significant effects on overall cognitive performance of age and education, but not for sex. In addition, we found that education predicts cognitive performance better than age.

Ageing leads to functional and structural brain deterioration, associated with cognitive decline and vulnerability to diseases such as dementia ^46^. Research in biological ageing has identified markers that recognise differences between individuals in their ageing rate and risk of cognitive decline ^47,48^. A meta-analysis revealed consistent evidence of declines in cognitive abilities such as memory, processing speed, and executive function with age ^49^. Furthermore, there is evidence of an indirect association of accuracy in working memory tasks and ageing ^50^. Our results align with the literature: MoCA performance decreases as age increases. However, many measures of biological age rely on associations with chronological age, which may not always be accurate ^51^.

Our observed effect of education on cognitive performance is consistent with the cognitive reserve theory ^52,53^. This view suggests that the educational level and other forms of cognitive stimulation can help offset the effects of ageing and brain pathology. A recent analysis of global longitudinal studies of cognitive ageing supports this theory by showing that education reduces the risk of the characteristic cognitive decline observed in dementia ^54^. Possibly, the development of cognitive skills due to formal education such as abstract reasoning, problem solving, language, and numerical literacy affects the observed performance on the MoCA. These results raise concerns related to the multiple challenges that living below the poverty line can create, such as malnutrition, that significantly affect educational achievement in Peru ^55^.

We only found sex differences in identification and attention, with men scoring higher than women. Current evidence supports sex differences in specific cognitive tasks, but not in overall cognitive performance: men generally perform better in mental rotation and visual–spatial working memory, while women show advantages in verbal and location memory ^56^. Similarly, there is evidence suggesting a faster decline in global cognition and executive function in women due to ageing ^57^. Our results may be compatible with this empirical evidence, showing that women score lower than men in identification and attention, possibly due to faster cognitive decline. However, our sample had an unequal sex distribution and the observed sex differences may be influenced by the cultural context ^58^.

We provide normative data and suggested cut-off scores for the MoCA adapted to the Peruvian population, fostering its clinical use and interpretation in the country. This pioneering study is the first to standardise the MoCA test in Peru, enriching its scientific landscape, and expanding its impact to other Spanish-speaking countries by setting a new benchmark for future studies focused on the cognitive decline of seniors ^24,42,59^.

One of the most notable findings of our study was the notably low score on the delayed recall test (semantic memory) and the high score on the (spatiotemporal) orientation test. The observed memory scores could be attributed to sample-specific impairments in the encoding, storage, and retrieval processes, or changes in the interaction between encoding and retrieval ^60,61^. In general, low delayed recall scores are associated with amnestic MCI ^62^ and may have negative implications on the daily activities of the evaluated individuals.

The exceptional performance in the orientation assessment indicates increased temporal and spatial acuity in our sample. This suggests preserved cognitive faculties and autonomy, relevant for excluding dementia diagnosis ^63^, as its loss indicates severe cognitive impairment in late-stage disease ^64^. The observed high orientation scores may be the result of cognitive reserve accumulated by work activity ^65^: Peruvians work more years on average than people in other countries and mainly under informal conditions ^66^. Thus, good orientation skills help maintain workplace competitiveness and secure basic economic subsistence. Future studies could explore the significance and social impacts of these findings.

To our knowledge, this study is the first post-pandemic standardisation of the test. This may explain the remarkably low scores observed in our sample compared to previous MoCA standardizations in Spanish-speaking countries ^42,59^. COVID-19 is thought to have direct impacts on cognitive functioning ^67,68^. Empirical evidence suggests that this viral infection produces changes in the orbitofrontal cortex and parahippocampal gyrus (decrease in gray matter), in the primary olfactory cortex (changes in markers of tissue damage) and a reduction in the overall size of the brain ^67^. Moreover, the implementation of social distancing protocols has had a profound impact on cognitive and physical activities, as well as social interactions. This disruption may have exacerbated levels of anxiety, depression, and stress responses ^69,70^, and has been associated with impairments in information processing speed and executive functions ^71^.

Hence, the results of our study are not directly comparable to any study carried out before the pandemic. We stress the need to update the original test standardisation to fit post-pandemic population characteristics. Adaptation to a post-pandemic environment may require adjustments to cognitive assessment expectations and methodologies. Such comprehensive revisions are critical to maintaining the relevance and accuracy of cognitive evaluations in this new era.

The study has limitations that must be taken into account when interpreting its results. Efforts to recruit a representative sample may still have under-represented certain demographics, affecting the external validity. Lack of long-term follow-up also limits the evaluation of the stability of the cognitive score and the prediction of future neurocognitive disorders. Another limitation is the lack of comparison with other cognitive evaluation tools and the absence of participants with a proper neuropsychological evaluation that confirms cognitive health. This affects the concurrent validity and sensitivity of the adapted MoCA compared to other instruments already available, limiting its use in the Peruvian context. Despite these limitations, the study represents an important step toward improving cognitive assessment in the elderly Peruvian population and highlights the need for more research in this field to address the cognitive health needs of the population in Latin America.

## Data Availability

All data produced in the present study are available upon reasonable request to the authors.

## Abbreviations

MoCA: Montreal Cognitive Assessment

## AUTHOR CONTRIBUTIONS

LB: Conceptualisation, Methodology, Investigation, Writing - Original Draft. EG: Conceptualisation, Methodology, Investigation, Writing - Original Draft. JV: Investigation, Writing - Original Draft. KM: Investigation, Writing - Reviewing and Editing. CM: Investigation, Writing - Reviewing and Editing. RP: Conceptualisation, Methodology, Software, Formal Analysis, Writing - Original Draft. SB: Supervision, Writing - Reviewing and Editing.

## ACKNOWLEDGMENTS

We acknowledge the research group Creativity, Technology and Talent (CREA TALENTUM), the Computational Cognitive Neuroscience Lab, and the Academic Direction of Social Responsibility (DARS) of the Pontifical Catholic University of Peru, for providing the resources required for this project. We also thank San José Obrero Polyclinic, San Miguel District Municipality and EDMECON, for facilitating access to participants. We thank Tiffany Sandoval for her support in the data collection process.

## ETHICAL STATEMENT

The ethical approval for the study was granted by the Ethics Committee for Research in Social Sciences, Humanities and Arts of the Pontifical Catholic University of Peru under approval number 145-2023-CEI-CCSSHHyAA. Participation in the study was voluntary and all participants gave their informed consent indicating their understanding of the objectives of the study and their willingness to participate.

## DATA SHARING AND DATA ACCESSIBILITY

The anonymised data sets and scripts employed for the analyses presented in this manuscript are accessible in the following public repository: https://github.com/neuropucp/MoCA-Peru

